# A Two-step Bayesian Mendelian Randomization Study on Cholecystitis and Dermatitis

**DOI:** 10.1101/2024.09.17.24313850

**Authors:** Chenyu Zhao, Changqian Cen, Ruihan Zhang, Wenjin He, Yiyang Jiao, Zhuoya Chen, Zhaoqi Wu, Ting Luan

**Affiliations:** Department of Critical Care Medicine, Shengjing Hospital of China Medical University, Shenyang, China; School of Pharmacy, Queen’s University Belfast, Belfast BT9 7BL, UK; School of Forensic Medicine, China Medical University, Shenyang, 110122, China; The First College of Clinical Medicine, Chongqing Medical University, Chongqing, China, 400016, China; School of Stomatology, China Medical University, Shenyang, 110122, China; Department of China Medical University-The Queen’s University of Belfast Joint College, School of Pharmacy, China Medical University, Shenyang, 110122, China

**Author notes:** Correspondence: Ting Luan. These authors contributed equally to this work (Chenyu Zhao, Changqian Cen).

**Keywords:** dermatitis, cholecystitis, mendelian randomization, bayesian weighted, IL-6, IL-7, IFN-gamma, HCG27, HLA-DRB5, SNP, GWAS

## Abstract

**Background:** Cholecystitis is an inflammatory disease involving the gallbladder, often associated with digestive disorders and systemic immune response. This systemic immune response could potentially influence the immune status of the skin, particularly in conditions like dermatitis. Despite extensive research on dermatitis, the causal relationship between cholecystitis and dermatitis subtypes (DSs) remains unclear.

**Objective:** The aim of this study was to investigate the causal relationship between cholecystitis and DSs.

**Methods:** Two-sample Mendelian randomization (MR) was used to analyze the causal relationship between cholecystitis and DSs. We then utilized the Bayesian Weighted Mendelian Randomization (BWMR) method to validate our findings and applied bidirectional MR analysis to confirm the causal direction. After establishing the associations between traits, we delved into the underlying mechanisms of this interesting finding. Subsequently, we used 91 inflammatory proteins as mediators and performed summary data-based mendelian randomization (SMR) analysis to further investigate the pathogenesis of DSs.

**Results:** MR results evidently showed that cholecystitis can significantly reduce the risk of allergic contact dermatitis (ACD) (IVW, OR=0.8834, p=0.0368) and exfoliative dermatitis (ED) (IVW, OR=0.5738, p=0.0126). BWMR also provided secondary validation of the casual associations. In the subsequent reverse direction MR analyses, reverse causality was not present, so cholecystitis had a unidirectional effect and existed as a protective factor for ACD and ED. Interestingly, cholecystitis appears to lower the risk of ACD and ED by downregulating IL-6, IL-7, and IFN-γ. Additionally, the genes HCG27 and HLA-DRB5 may play a significant role in the pathogenesis of ACD.

**Conclusion:** This study used a two-step MR analysis of genetic summary data to investigate to what extent inflammatory proteins impact the protective role of cholecystitis on dermatitis. We also identified several proteins and genes that could serve as potential drug targets.

*Graphical abstract:* 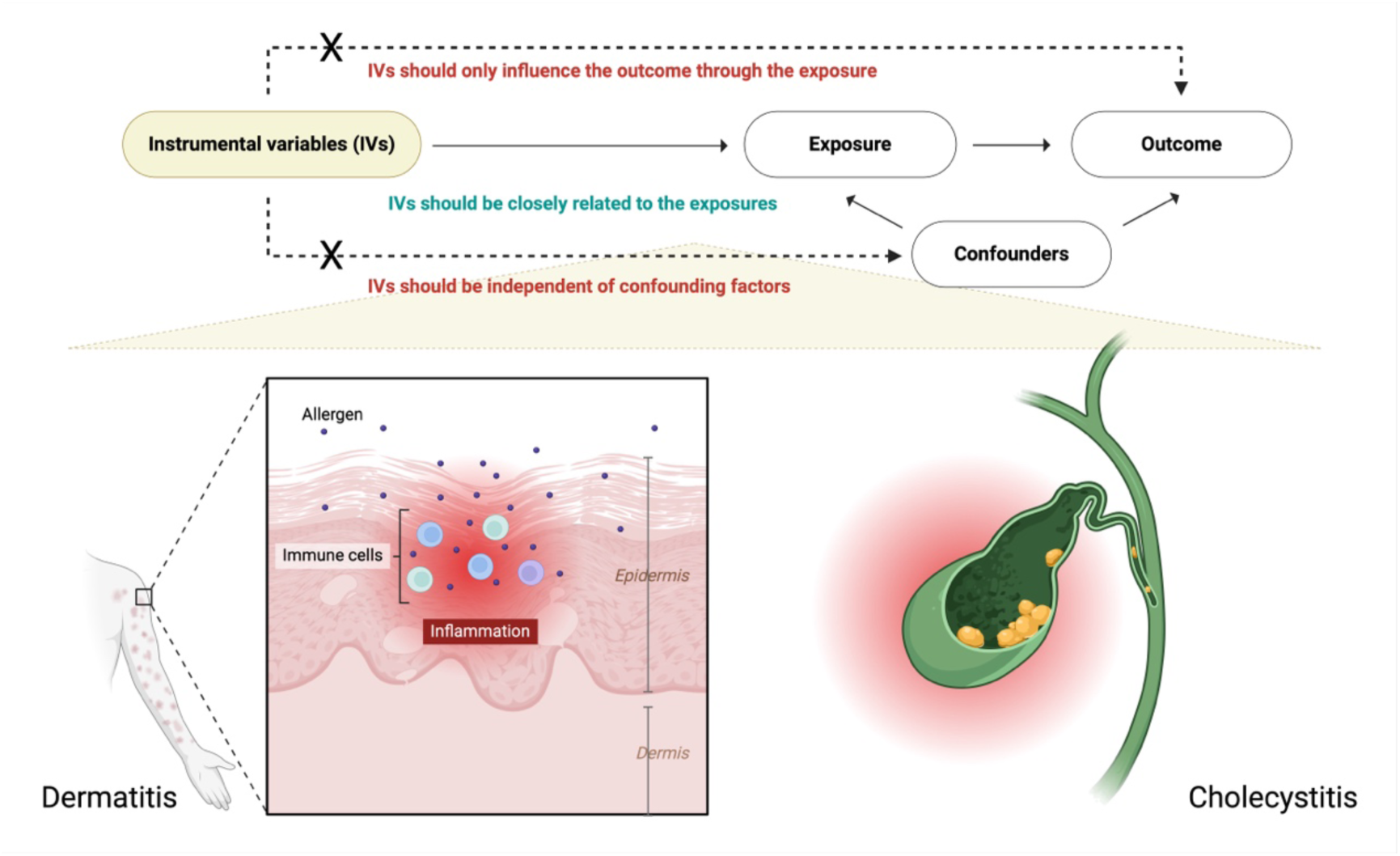

## Introduction

Dermatitis, a prevalent inflammatory condition of the skin, presents significant clinical challenges due to its diverse symptomatology, including redness, swelling, dryness, itching, blisters, scabbing, and peeling [1]. The primary subtypes of dermatitis—contact dermatitis, atopic dermatitis, and seborrheic dermatitis—are well-known for their chronic and often debilitating nature, affecting patients’ quality of life [2][3][4]. Despite ongoing advancements in dermatological research, the underlying causes and risk factors for different dermatitis subtypes (DSs) remain only partially understood, leaving gaps in effective prevention and treatment strategies [5][6].

Allergic contact dermatitis (ACD) is an immune-mediated inflammatory reaction triggered by skin contact with allergens [7]. ACD typically presents with erythema, pruritus, vesicles, and scaling. Common triggers include metals (such as nickel), cosmetics, preservatives, and plants [8]. Upon contact with an allergen, T cells in the immune system recognize and overreact to these substances, leading to inflammation and the characteristic symptoms of dermatitis. Exfoliative dermatitis (ED), also known as erythroderma, is a severe and widespread inflammatory condition of the skin characterized by extensive erythema, scaling, and shedding of the skin [9]. ED is often caused by adverse drug reactions, exacerbation of chronic skin conditions such as psoriasis or eczema, or certain systemic diseases [6][10]. It is frequently associated with systemic symptoms like fever, lymphadenopathy, and fluid loss, and in severe cases, it can be life-threatening. Cholecystitis, an inflammation of the gallbladder, is typically associated with conditions like gallstones and bile duct obstruction [11]. However, recent studies have suggested that its impact may extend beyond the biliary system. Cholecystitis has been shown to influence systemic inflammation, potentially modulating immune responses in ways that could affect other inflammatory diseases [12]. The exact mechanisms through which cholecystitis might exert protective effects on conditions like dermatitis are not yet fully elucidated, warranting further investigation.

Mendelian randomization (MR) is an increasingly utilized method in epidemiology that leverages genetic variants as instruments to assess causal relationships between risk factors and diseases [13]. We initially conducted two-sample MR analysis method using cholecystitis as the exposure to evaluate its impact on various DSs. For DSs, we introduced the innovative use of Bayesian weighted mendelian randomization (BWMR) as a secondary validation method for phenotype associations. BWMR enhances precision and robustness, especially in studies with complex genetic architectures or when genetic instruments vary in strength [14]. We then applied bidirectional Mendelian Randomization and two-step Mendelian Randomization to confirm the causal direction and assess the influence of inflammatory protein mediators [15]. Finally, we performed the summary data-based mendelian randomization (SMR) analysis to further investigate the drug targets of ACD and ED [16].

This study aims to explore the genetic relationship between cholecystitis and dermatitis subtypes using a two-step MR approach. By analyzing data from genome-wide association studies (GWAS), we sought to determine whether cholecystitis serves as a protective factor against specific forms of dermatitis and to identify the genetic and molecular mechanisms underlying this potential relationship.

## Method

### Study design

MR, a specialized analysis that utilizes genetic instrumental variables (IVs), employs single nucleotide polymorphisms (SNPs) as IVs to assess the impact of risk factors on various outcomes, particularly disease. SNPs identified in GWAS as being linked to specific risk factors can serve as IVs to test their causal effects on different outcomes. The overall study is progressively advancing from disease phenotypes to molecular mechanisms, with the primary aim of identifying new treatment approaches for dermatitis. The selection of IVs dwelled upon three pivotal assumptions: (1) the IVs must be significantly connected to the exposure; (2) the IVs cannot be connected to any known confounders that could alter the association between an exposure and an outcome; and (3) the IVs must be unrelated to the outcomes and may only affect the outcomes through their effects on the exposure [17].

Two-sample MR was employed to explore the causal relationship between cholecystitis and DSs initially. To ensure the robustness of our findings, we utilized the BWMR method and conducted bidirectional MR analysis to confirm the causal direction of the observed associations. After establishing the relationship between these traits, we investigated the underlying mechanisms behind this intriguing connection [18]. To do this, we conducted Gene Ontology (GO) and Kyoto Encyclopedia of Genes and Genomes (KEGG) enrichment analyses on SNPs strongly associated with cholecystitis, which highlighted the potential involvement of inflammatory proteins. Building on these insights, we selected 91 inflammatory proteins as potential mediators and performed SMR analysis to further investigate the pathogenesis of allergic contact dermatitis ACD and ED [19][20]. These analyses allowed us to delve deeper into the biological pathways and potential molecular mechanisms linking cholecystitis to these skin conditions (Figure 1).

**Figure 1.**
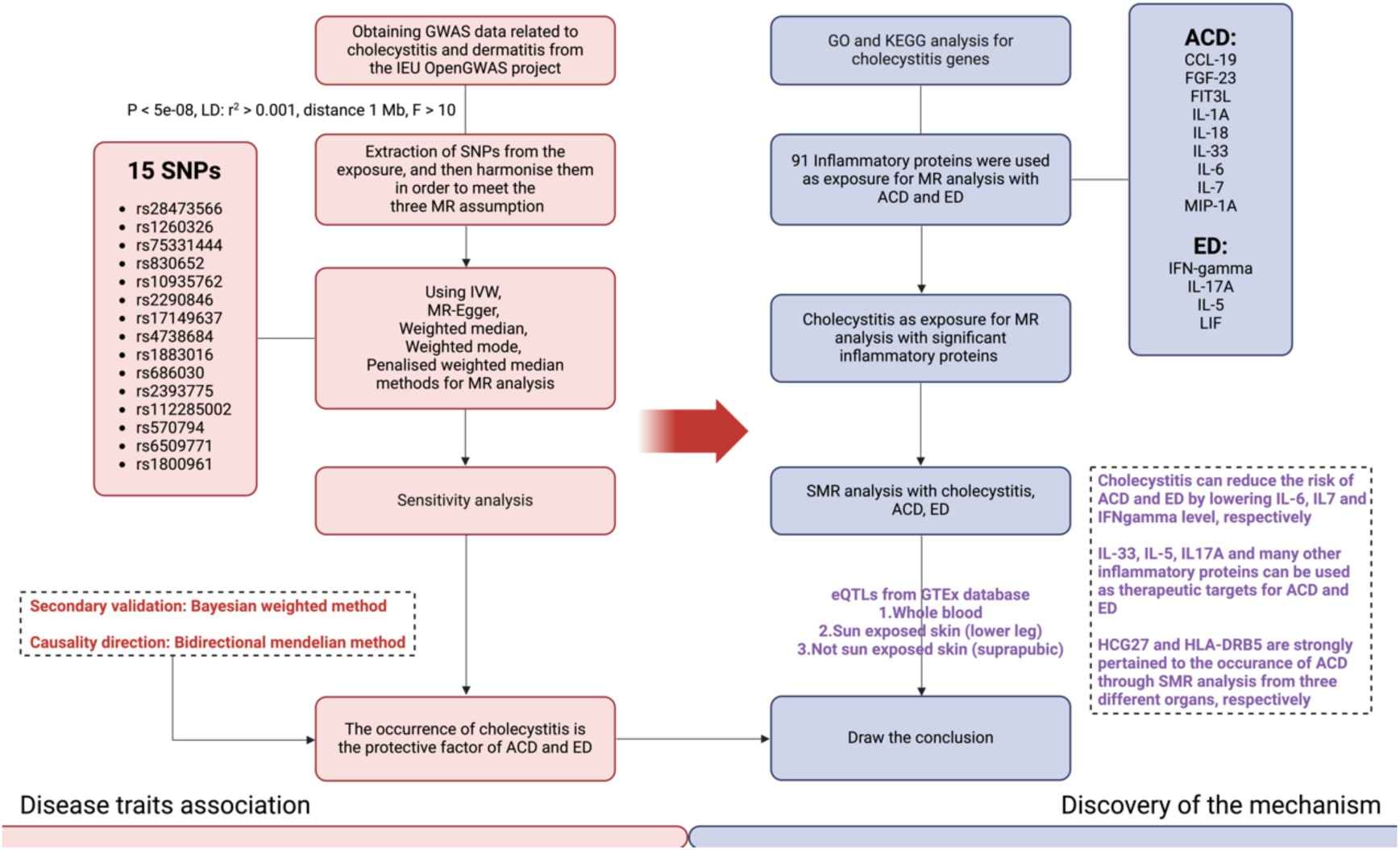
The overall study is progressively advancing from disease phenotypes to molecular mechanisms, with the primary goal of identifying new treatment strategies for dermatitis. The research can be broadly divided into two parts: first, establishing causal relationships, and then uncovering the underlying mechanisms.

### Data Source

In the initial two-sample Mendelian analysis, exposure and outcome data were obtained from the IEU OpenGWAS project and FinnGen, respectively (Supplementary Table 1).

The 91 inflammatory proteins were sourced from the GWAS Catalog. For all MR analyses, we utilized GWAS data from different consortiums to prevent sample overlap and potential bias in the results. Additionally, we restricted the population to individuals of European descent, thereby minimizing the influence of population-specific genetic factors. For the SMR analysis, we downloaded expression quantitative trait locus (eQTL) data from three different organs from the GTEx database for analysis. All data sources are detailed at the end of the article [21].

### IVs selection

The selection of genetic instruments adhered to the following criteria: First, SNPs were included if they showed a significant genome-wide association with the exposure (p < 5 × 10−8 for cholecystitis; p < 1 × 10−5 for inflammatory proteins, ACD, and ED). We applied pairwise linkage disequilibrium (LD) thresholds from the original GWAS for each mediating trait, with SNPs for each trait adhering to an LD cut-off of r2 < 0.001 within a window of 1 MB. The strength of the included SNPs was assessed with F-statistics. Weak instrument with a F-statistic less than 10 would be excluded from the IVs. For the qualified IVs, we conducted gene annotation using resources from the National Center for Biotechnology Information (NCBI) (Supplementary Table 2).

### Statistical analysis of MR

MR analyses were conducted using the ‘TwoSampleMR’ package in R (version 4.2.3, http:// www.R-project.org, The R Foundation). We first applied the Inverse-Variance Weighted (IVW) method to combine the genetic instruments strongly associated with cholecystitis in order to estimate its causal relationship with various DSs. Additionally, we employed methods such as MR Egger, Weighted median, Weighted mode and Penalized weighted median to provide more evidence of our findings. For exposures and outcomes where the IVW p-value < 0.05, we then innovatively used the BWMR method for a second round of validation to enhance the reliability of the causal relationship. BWMR, a MR method based on Bayesian inference, models the prior distribution of instrument variable effects to provide a more robust causal effect estimate. This method is particularly effective at addressing potential horizontal pleiotropy and offers reliable estimates when instrument variable effects are imbalanced. MR also provides strong directionality validation; for positive results obtained from the above methods, we performed bidirectional MR testing, with the IVW results serving as the primary standard. Finally, we conducted a two-step MR analysis. Given that cholecystitis often leads to immune system dysregulation, we selected inflammatory proteins within the body as mediators. In the two-step MR, we first treated inflammatory proteins as exposures and conducted MR analyses with the previously identified positive DSs. For the positive results, we performed further analysis by treating cholecystitis as the exposure and conducting MR analyses with the inflammatory proteins. We defined the β value from the MR analysis between cholecystitis and dermatitis as β0. In the two-step analysis involving inflammatory proteins, the beta value with cholecystitis as the exposure was labeled as β1, and the β value with dermatitis as the outcome was labeled as β2. The mediation effect is calculated by multiplying the effect estimates from the first and second steps, that is, mediation effect = β1 × β2. The direct effect is then determined by subtracting the mediation effect from the total effect, that is, direct effect = β0 - β1 × β2. The mediation proportions were calculated according to the formula: (β1 × β2)/β0. Results were presented as an odds ratio (OR) along with a 95% confidence interval (CI) per standard deviation. Standard errors and CIs were calculated using delta methods.

For all the MR analysis results mentioned above, we conducted sensitivity tests, including heterogeneity tests, pleiotropy tests, and leave-one-out analysis to ensure the robustness and reliability of our findings. The heterogeneity test, using Cochran’s Q statistic, assessed whether multiple IVs consistently estimated the causal effect, with a p-value > 0.05 indicating no significant heterogeneity among the IVs. The pleiotropy test evaluated whether IVs influenced the outcome through multiple biological pathways rather than solely through the exposure. P-value > 0.05 suggested the absence of horizontal pleiotropy; if pleiotropy was detected, the MR-PRESSO method was employed to remove outliers and correct the results. Additionally, leave-one-out analysis was used to assess the impact of individual IVs on the overall causal estimate by sequentially excluding each IV and observing any significant changes in the estimate, helping identify SNPs with a major influence and ensuring the stability of the findings.

### SMR analysis

SMR is a MR method that integrates gene expression data with GWAS data to evaluate potential causal relationships between gene expression and phenotypes, such as diseases. The core principle of SMR is to use genetic variants associated with gene expression, known as eQTLs, as IV to infer causal links between gene expression and disease outcomes. In our analysis, we selected three eQTL datasets from the GTEx database and proceeded through several key steps: SNPs significantly associated with specific gene expressions as IV, using MR methods to estimate the causal effects of these gene expressions on diseases or phenotypes, and performing the heterogeneity in dependent instruments test (HEIDI) to detect horizontal pleiotropy, thereby distinguishing true causal effects from false positives. We identified significant genes by filtering for instrumental variables with a p-value < 0.05 and a HEIDI test p-value > 0.05. SMR analyses were then conducted for cholecystitis, DSs individually, followed by identifying overlapping results between cholecystitis and DSs.

## Results

### Causal association of cholecystitis and DSs

In the initial two-sample MR analysis, we identified a total of 15 IVs that could serve as proxies for cholecystitis. Upon closely examining the IVW results, it was unexpectedly revealed that a causal relationship exists between cholecystitis and both ACD and ED, where cholecystitis appears to act as a protective factor. Results suggest that cholecystitis may have a significant protective effect against certain DSs, specifically ACD and ED (Figure 2) (Supplementary Figures 1-9). For ACD, the IVW method indicates a statistically significant association with a p-value of 0.0368 and an OR of 0.8834, suggesting that cholecystitis could reduce the risk of developing this condition. This finding is consistent across multiple methods, reinforcing its robustness. Similarly, for ED, the IVW method reveals a significant association with a p-value of 0.0045 and an OR of 0.5738, indicating that cholecystitis might lower the likelihood of it (Figure 3). In sensitivity analysis, we did not observe any evidence of bias (Table 1)(Figure 4).

**Figure 2.**
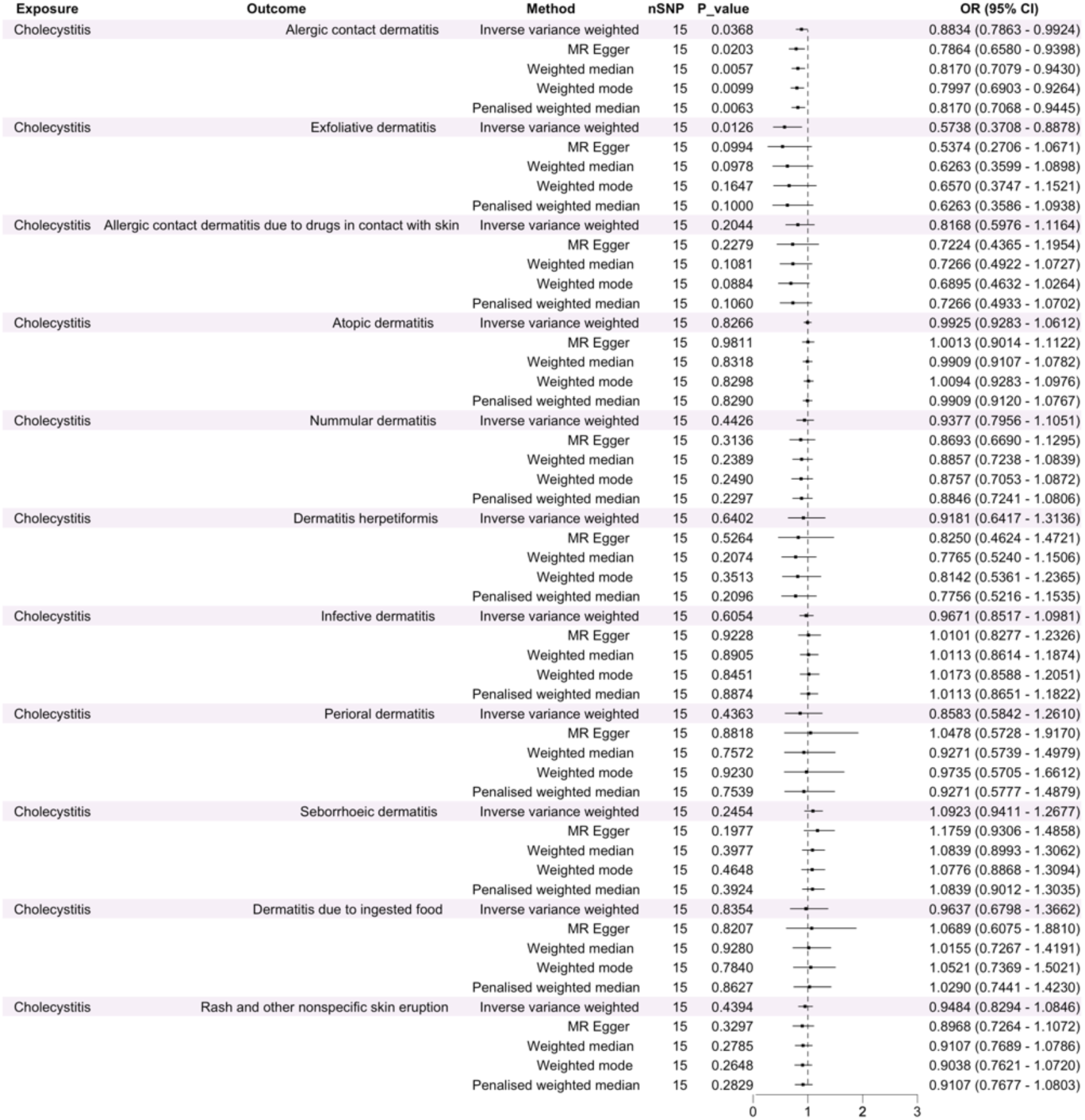
Forest plot of MR analysis with cholecystitis as the exposure and different DSs as the outcomes.

**Figure 3.**
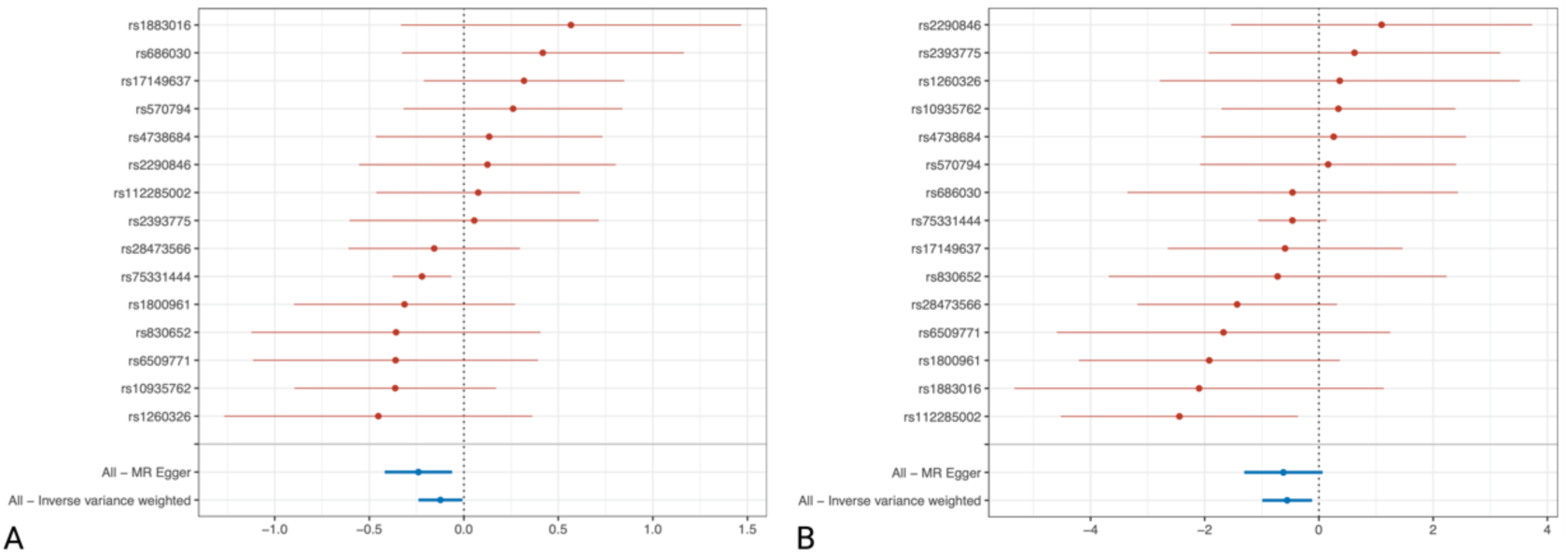
A. Forest plot of MR analysis with cholecystitis as the exposure and ACD as the outcome. B. Forest plot of MR analysis with cholecystitis as the exposure and ED as the outcome.

**Figure 4.**
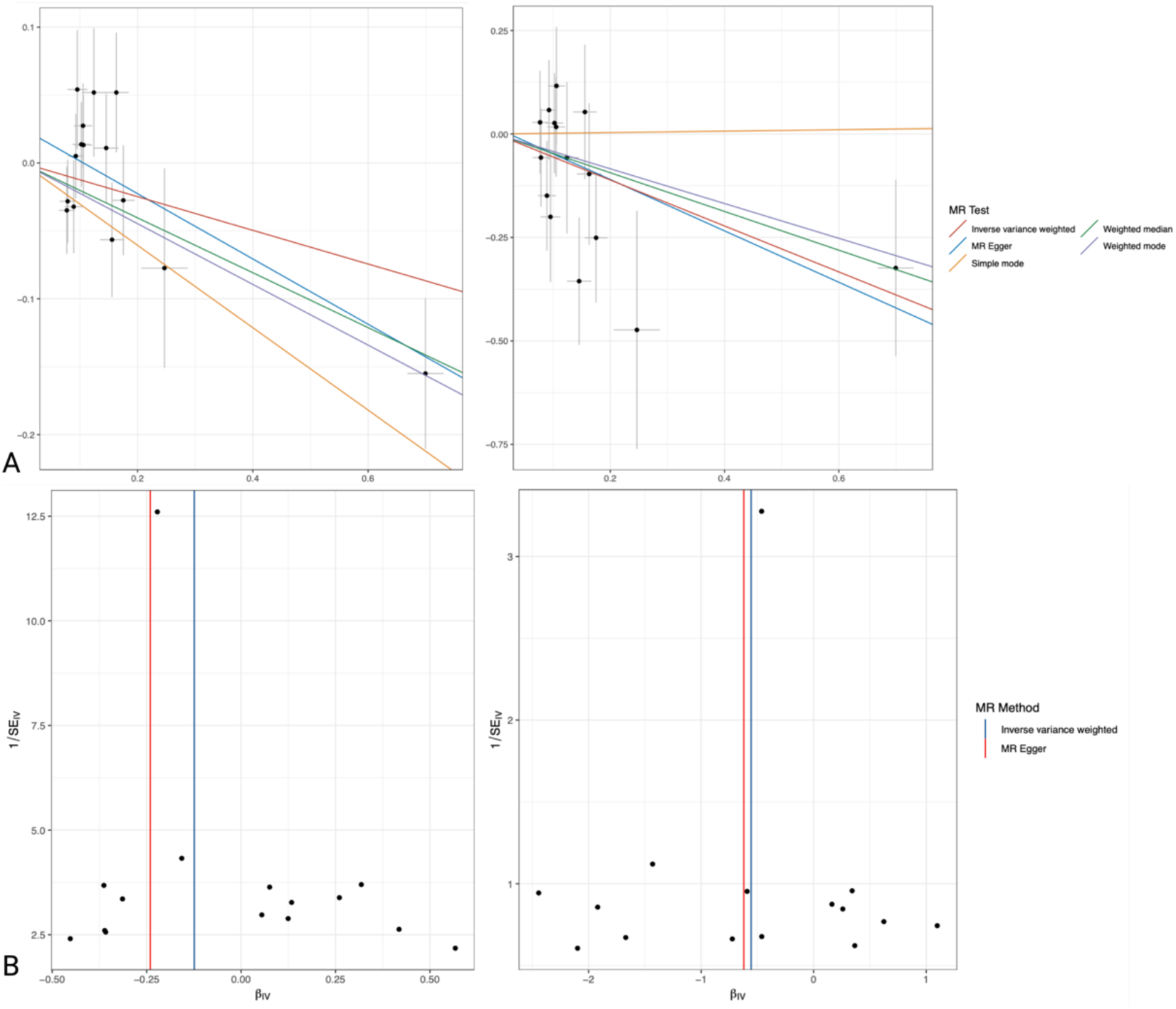
A. Scatter plot and funnel plot of MR analysis with cholecystitis as the exposure and ACD as the outcome. B. Scatter plot and funnel plot of MR analysis with cholecystitis as the exposure and ED as the outcome.

**Table 1.** Cholecystitis as exposure to MR Analysis sensitivity test results.

| **Exposure** | **Outcome** | **Egger_intercept** | **SE** | **Pleiotropy_pval** |
| --- | --- | --- | --- | --- |
| Cholecystitis | Allergic contact dermatitis | 0.0255 | 0.0154 | 0.1213 |
|  | Exfoliative dermatitis | 0.0144 | 0.0591 | 0.8120 |
| **Exposure** | **Outcome** | **Method** | **Q** | **Q_pval** |
| Cholecystitis | Allergic contact dermatitis | Inverse variance weighted | 14.7774 | 0.3935 |
|  |  | MR Egger | 12.0299 | 0.5252 |
|  | Exfoliative dermatitis | Inverse variance weighted | 11.3145 | 0.6612 |
|  |  | MR Egger | 11.2556 | 0.5894 |

In contrast, for the other DSs analyzed—such as atopic dermatitis, nummular dermatitis, and seborrheic dermatitis—the p-values are generally above 0.05, indicating no significant causal relationship with cholecystitis. These results suggest that while cholecystitis might be linked to a reduced risk of specific DS, its influence on other forms of dermatitis appears to be minimal or non-significant.

### BWMR validation for positive casual association

Subsequently, we treated cholecystitis as an exposure and conducted BWMR analysis on both ACD and ED. For ACD, the OR was 0.8825 with a p-value of 0.033. Similarly, for ED, we obtained an OR of 0.5707 with a p-value of 0.0136. These results suggest that cholecystitis indeed serves as a protective factor against both ACD and ED.

### Bidirectional and two-step MR analysis

In the reverse MR analysis, negative results were consistently observed in the IVW method, leading us to conclude that cholecystitis has a unidirectional effect on ACD and ED (Figure 5C).

**Figure 5.**
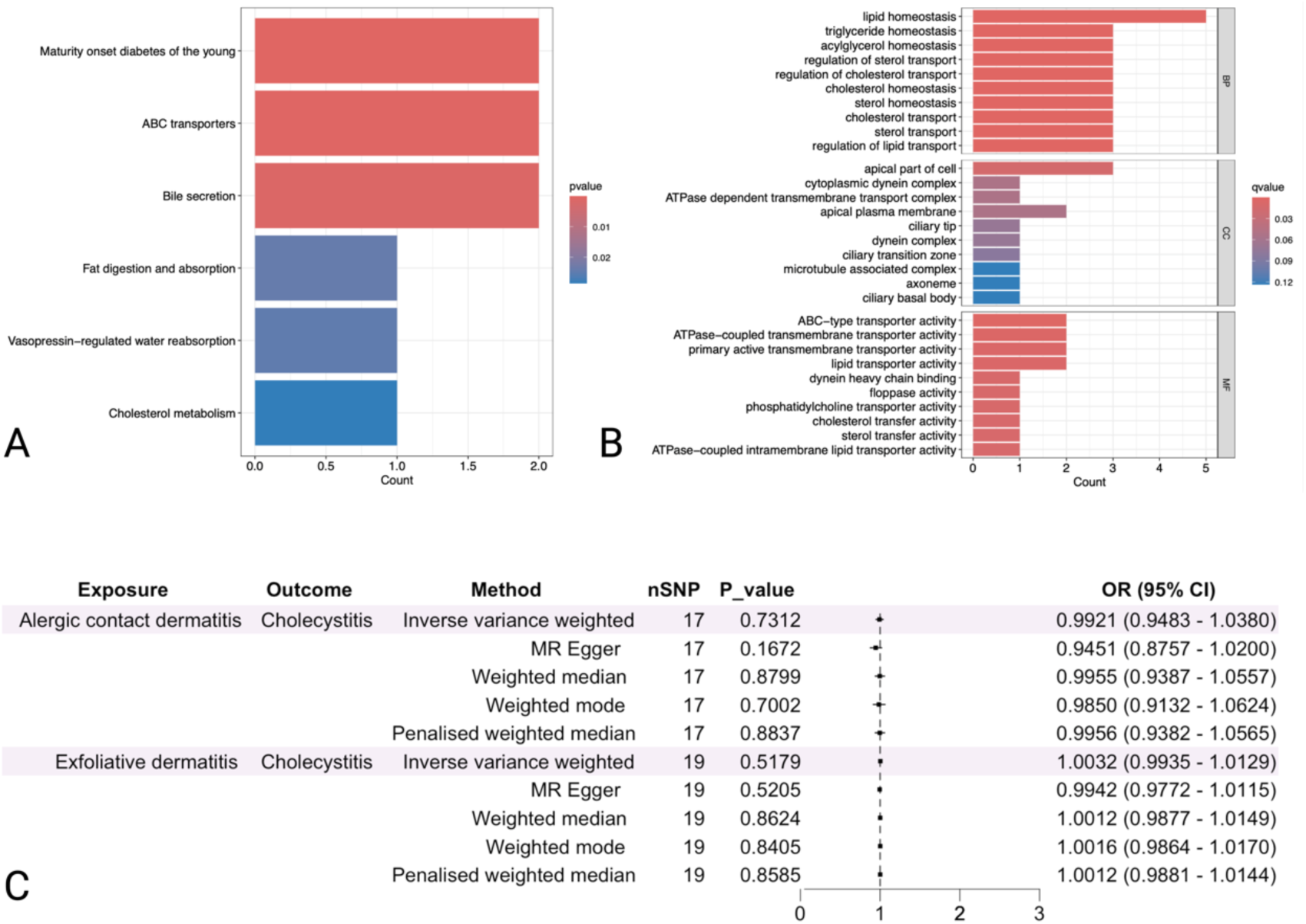
A. KEGG enrichment analysis of annotated genes of IVs. B. GO analysis of annotated genes of IVs. C. Reverse direction MR analysis with ACD and ED as the exposures.

By annotating the IVs significantly associated with cholecystitis and performing GO and KEGG enrichment analyses, we propose that cholecystitis may primarily exert its effects on ACD and ED through inflammation-related functions and pathways. Therefore, we selected 91 inflammation-related proteins for further MR analysis (Supplementary Tables 3-5).

We identified nine inflammation proteins with a causal relationship to ACD, among which C-C motif chemokine (CCL-19) and interleukin-1alpha (IL-1A) levels were inversely associated with the occurrence of ACD. Notably, IL-33, IL-6, and IL-7 had a more significant impact on ACD, with ORs exceeding 1.2, warranting our focused attention. Additionally, four inflammation proteins demonstrated a causal relationship with ED; aside from leukemia inhibitory factor (LIF), the remaining proteins were identified as risk factors. The results related to inflammation proteins associated with ED were highly significant (Figure 6A).

**Figure 6.**
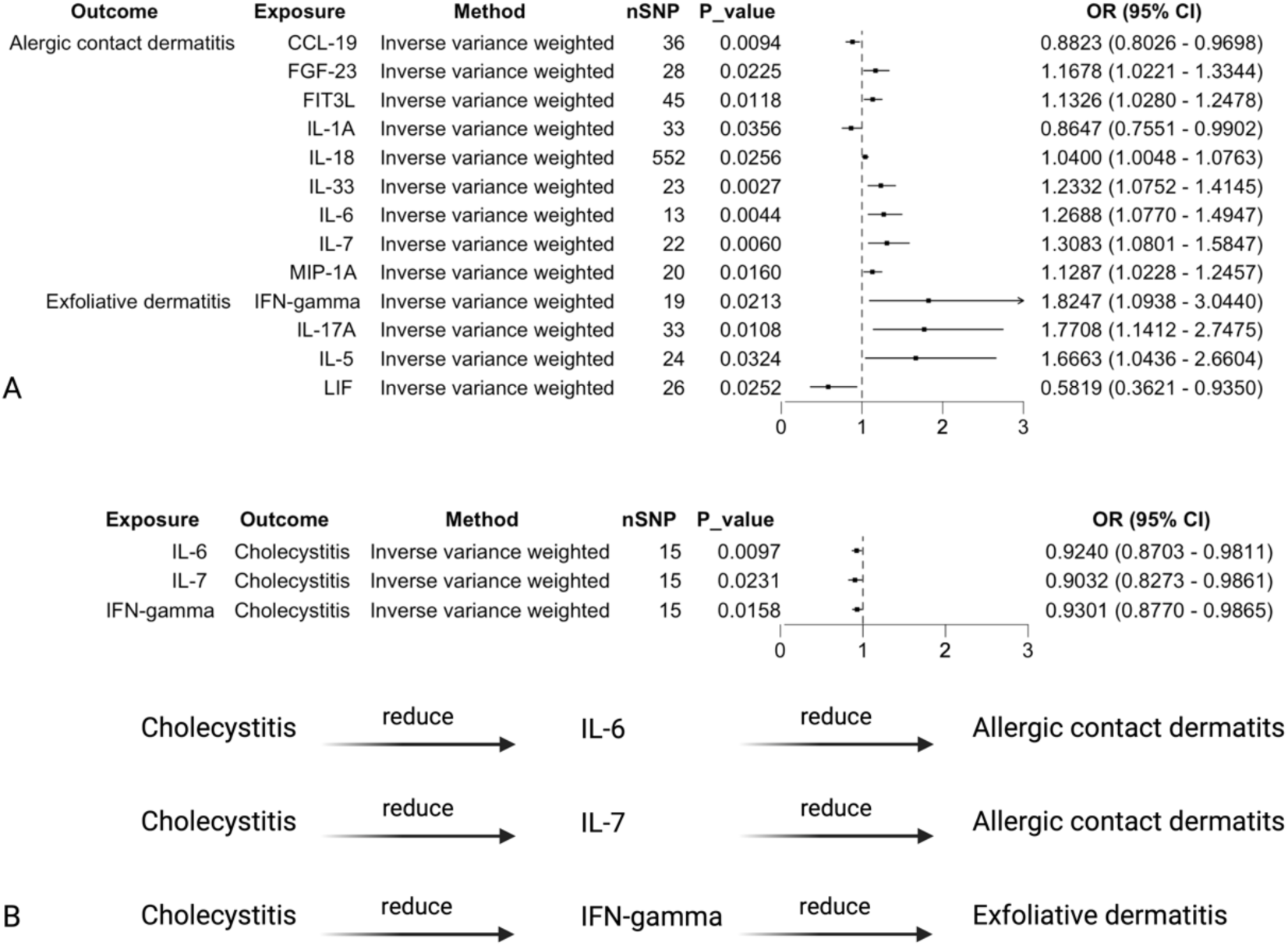
Two-step MR with inflammatory as the mediator. A. MR analysis with inflammatory proteins as the exposure. B. MR analysis with inflammatory proteins as the outcome.

In the MR analysis, where inflammation proteins were treated as outcomes, we found that cholecystitis likely exerts a protective effect against ACD through IL-6 and IL-7 as intermediary inflammation proteins. Similarly, cholecystitis appears to offer protective effects against ED via interferon gamma (IFN-γ) (Figure 6B).

### SMR analysis

In the SMR results, no significant genetic associations were identified between cholecystitis and ED across three eQTL datasets. However, in the analysis for ACD, two critical genes, histocompatibility antigen-DRB5 (HLA-DRB5) and HLA Complex Group 27 (HCG27) were identified. These genes were consistently present in whole blood and two skin tissue datasets (Figure 7A). Given their consistent results across the three eQTL datasets, we consider these genes to be potential therapeutic targets for treating ACD (Figure 7B).

**Figure 7.**
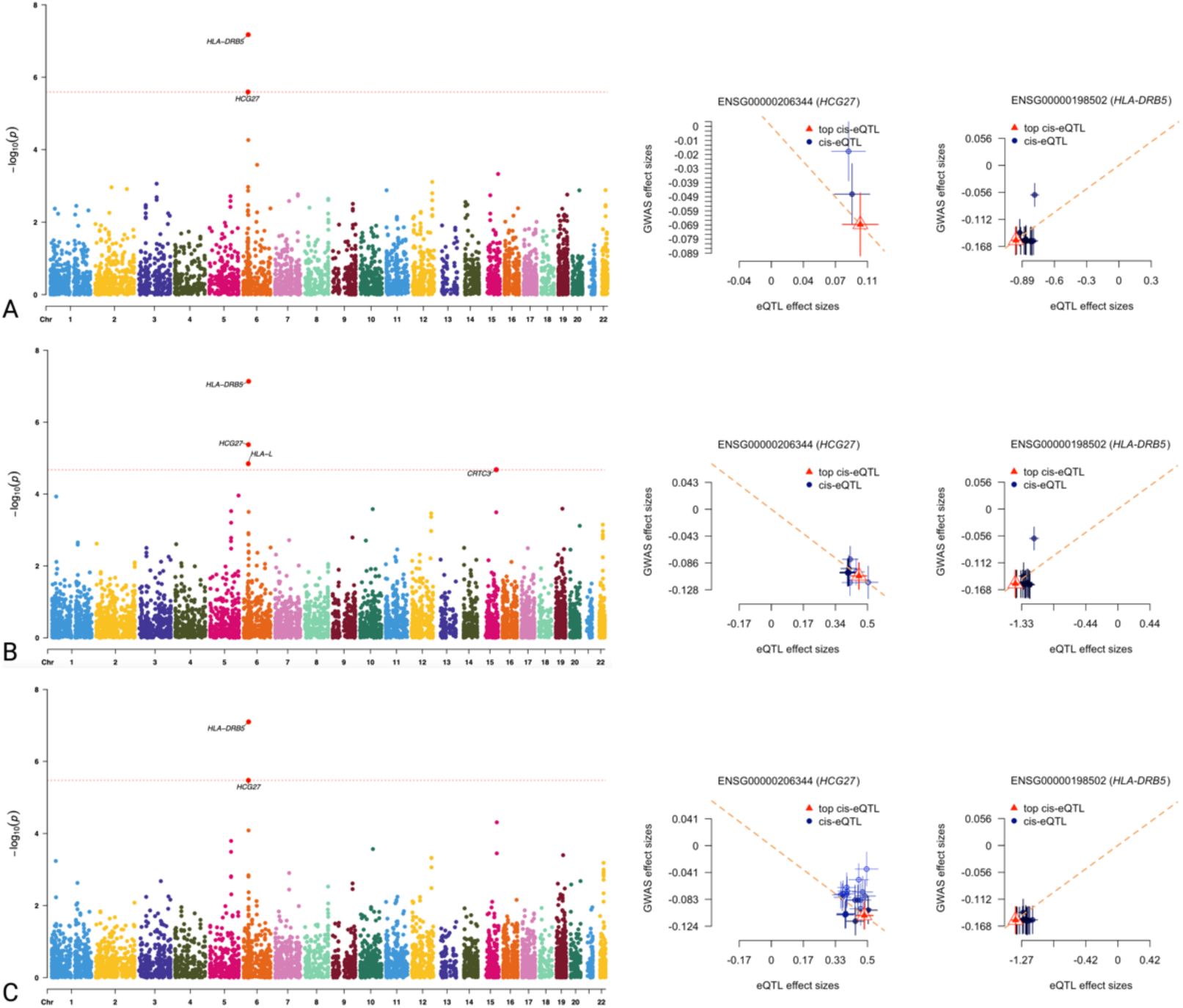
SMR analysis for ACD. A. Manhattan plot and single-line plot using whole-blood eQTL data as the IVs. B. Manhattan plot and single-line plot using sun exposed lower leg eQTL data as the IVs. C. Manhattan plot and single-line plot using not sun exposed suprapubic eQTL data as the IVs.

## Discussion

Our work yielded substantial findings, exploring the potential connections between cholecystitis and different DSs, with an emphasis on identifying biomarkers for treating ACD and ED. Initially, we were quite surprised to observe that cholecystitis might act as a protective factor. Although we found positive MR causal associations only in ACD and ED, the results from other DSs are also noteworthy.

The "gut-skin axis" concept has received some scientific support, though its specific mechanisms remain under investigation [22][23][24]. Moreover, we have previously observed the concurrent occurrence of calculous cholecystitis and skin rashes in several cases [25][26][27]. Given the gallbladder’s critical role in the digestive system, the mechanisms by which it might reduce the incidence of dermatitis are certainly worth exploring. Based on clinical knowledge and previous research, we hypothesize several mechanisms. First, the immune response triggered by cholecystitis might enhance systemic immune surveillance, potentially reducing skin inflammation [28]. Secondly, alterations in bile acid metabolism may influence the gut microbiota, indirectly affecting skin immune responses. Additionally, impaired gallbladder function could lead to decreased absorption of fats and fat-soluble vitamins, which are closely linked to skin health. This reduced absorption might alter the mechanisms of skin inflammation [29][30].

Fortunately, in our two-step MR analysis, we identified that IL-6, IL-7, and IFN-gamma play mediating roles in the immune response. Several studies have shown elevated serum levels of pro-inflammatory cytokines (IL-1α, IL-6, IL-7) in patients with gallstones [31]. Specifically, IL-6 is strongly positively correlated with both acute and chronic cholecystitis [32]. Research by Yulu Wang and colleagues highlighted that extracellular vesicle from Clostridium, which is strongly associated with cholecystitis, can activate Toll-like receptor 9 and promote the expression of IL-6 and tumor necrosis factor alpha (TNF-α) in mouse biliary epithelial cells [33]. Importantly, IL-6 is also deeply implicated in dermatitis; IL-6 deficiency exacerbates skin inflammation in mouse models of irritant dermatitis, likely due to IL-6’s negative regulation of IL-22Rα expression on epidermal keratinocytes, thereby reducing inflammation [34] [35][36]. In contrast, the relationship between cholecystitis and IL-7 levels has been less studied, though immunohistochemical staining in 20 children with calculous cholecystitis did show some differences [37]. Our results indicate that IL-7 has a significant impact on ACD, although most current research links it primarily with atopic dermatitis. Nevertheless, these findings provide some useful insights[38][39][40]. Additionally, a decrease in IFN-γ levels has been observed after routine cholecystectomy, underscoring the gallbladder’s important role in regulating IFN-γ levels in the body [41]. ACD is an inflammatory type IV hypersensitivity disorder, characterized by skin infiltration induced by polyclonal effector CD8 αβ T cells and tissue-resident memory T cell precursors. Paeoniflorin has been shown to possess anti-allergic and anti-inflammatory properties in treating ACD [42]. Xiaoting Wu et al. demonstrated that Paeoniflorin could also mitigate ACD responses in a mouse model by inhibiting the production of IFN-γ in T lymphocytes [43]. Overall, our findings are largely consistent with previous research.

Furthermore, we aimed to perform additional analyses using RNAseq data from the GEO database. Unfortunately, we were unable to find suitable datasets for cholecystitis, ACD, or ED. To identify core pathogenic genes associated with these diseases, we considered using the SMR approach, intending to obtain individual results for each disease and then identify common genes by intersecting these results. However, we were only able to identify positive genes in ACD. In the SMR analysis of ACD, we discovered that the eQTL data from three different tissues consistently highlighted two genes: HCG27 and HLA-DRB5. Both HCG27 and HLA-DRB5 are located in the MHC region of chromosome 6, an area known to contain numerous genes related to immune response and autoimmune diseases. Given their proximity in the same genetic region, it is possible that they share expression patterns or regulatory mechanisms, potentially working together in the immune system to influence the strength and nature of immune responses. Although the direct functional relationship between these two genes remains unclear, their co-expression in certain immune-related diseases suggests they may be involved in shared immune pathways. Current research indicates that the role of HCG27 in dermatological diseases remains unknown. In contrast, HLA-DRB5 has been identified as a pathogenic gene in various skin disorders. Further studies are needed to clarify the functions of these two genes.

## Conclusion

In conclusion, this study advances our understanding of the complex interactions between cholecystitis and skin inflammation, particularly in the context of ACD and ED. The findings suggest that cholecystitis, through its modulation of inflammatory responses, could serve as a protective factor against certain forms of dermatitis. This novel insight opens up potential avenues for developing new therapeutic strategies that leverage this protective effect.

Further research is warranted to explore the broader implications of these findings and to determine whether similar protective mechanisms might exist for other inflammatory skin conditions. Additionally, the identification of specific inflammatory proteins and genetic markers provides a foundation for future studies aimed at developing targeted treatments for DSs.

## Data Availability

All data produced in the present study are available upon reasonable request to the authors

https://gwas.mrcieu.ac.uk/

https://www.ebi.ac.uk/gwas/

## Abbreviation

DSs: Dermatitis subtypes
MR: Mendelian randomization
BWMR: Bayesian weighted mendelian randomization
ACD: Allergic contact dermatitis
ED: Exfoliative dermatitis
SMR: Summary data-based mendelian randomization
GWAS: Genome-wide association studies
IV: Instrumental variable
SNP: Single nucleotide polymorphism
GO: Gene Ontology
KEGG: Kyoto Encyclopedia of Genes and Genomes
eQTL: Expression quantitative trait locus
LD: Linkage disequilibrium
NCBI: National Center for Biotechnology Information
IVW: Inverse variance weighted
OR: Odds ratio
CI: Confidence interval
HEIDI: Heterogeneity in dependent instruments test
CCL19: C-C motif chemokine 19
FGF-23: Fibroblast growth factor 23
Flt3L: Fms- like tyrosine kinase 3
IL: Interleukin
MIP-1A: Macrophage inflammatory protein 1a
IFN-γ: Interferon gamma
LIF: Leukemia inhibitory factor
TNF-α: Tumor necrosis factor alpha
HLA-DRB5: Histocompatibility antigen-DRB5
HCG27: HLA Complex Group 27
MHC: Major histocompatibility complex

## Data availability

Cholecystitis and all DSs GWAS data are available through the MRC IEU Open GWAS database (https://gwas.mrcieu.ac.uk/).

All inflammatory protein GWAS data are available through the GWAS Catalog (GCST90274758-GCST90274848) (https://www.ebi.ac.uk/gwas/).

The SMR software and eQTL data are available from Yang Lab of Westlake University (https://yanglab.westlake.edu.cn/software/smr/#Overview).

## Author Contribution Statement

Chenyu Zhao and Changqian Cen contributed equally to this work. Chenyu Zhao conceptualized and drafted the manuscript. Changqian Cen designed the figures. Wenjin He and Ruihan Zhang provided the information of the manuscript. Yiyang Jiao and Zhuoya Chen edited the manuscript. Zhaoqi Wu was responsible for language check. Ting Luan revised the manuscript. All authors read and approved the final manuscript.

## Acknowledgements

We express our deep gratitude to all the researchers working in the field of treating dermatitis. At the same time, we thank China Medical University for being such a superior platform to provide a bridge for clinicians and researchers in affiliated institutions to help each other.

## Ethical Statement

There is no human or animal studies involved in this work.

