## Supplementary Table 1 for "A Two-step Bayesian Mendelian Randomization Study on Cholecystitis and Dermatitis"

| **Exposure** | **Exposure ID** | **Sample size** | **Case/Control** | **Population** | **SNP number** | **Year** |
| --- | --- | --- | --- | --- | --- | --- |
| Cholecystitis | ebi-a-GCST90018818 | 471,251 | 9,820/461,431 | European | 24,183,039 | 2021 |
| **Outcome** | **Outcome ID** | **Sample size** | **Case/Control** | **Population** | **SNP number** | **Year** |
| Allergic contact dermatitis | finn-b-L12_ALLERGICCONTACT | 200,944 | 2,204/198,740 | European | 16,380,431 | 2021 |
| Allergic contact dermatitis due to drugs in contact with skin | finn-b-ALLERGIC_CONTACT_DERMA_DRUGS_CONTACT_W_SKIN | 218,792 | 296/218,496 | European | 16,380,466 | 2021 |
| Atopic dermatitis | finn-b-L12_ATOPIC | 205,764 | 7,024/198,740 | European | 16,380,443 | 2021 |
| Dermatitis herpetiformis | finn-b-L12_DERMATHERP | 218,344 | 278/218,066 | European | 16,380,466 | 2021 |
| Exfoliative dermatitis | finn-b-L12_EXFOLIATIVE | 198,883 | 143/198,740 | European | 16,380,424 | 2021 |
| Infective dermatitis | finn-b-L12_INFECT_DERM | 200,460 | 1,720/198,740 | European | 16,380,426 | 2021 |
| Perioral dermatitis | finn-b-L12_PERIORALDERM | 211,324 | 185/211,139 | European | 16,380,451 | 2021 |
| Seborrhoeic dermatitis | finn-b-L12_SEBORRHOEIC | 199,993 | 1,253/198,740 | European | 16,380,425 | 2021 |
| Nummular dermatitis | finn-b-L12_DERM_NUMMULAR | 199,796 | 1,056/198,740 | European | 16,380,428 | 2021 |
| Rash and other nonspecific skin eruption | finn-b-R18_RASH_OTHER_NONSPECIFIC_SKIN_ERUPT | 209,088 | 1,540/207,548 | European | 16,380,436 | 2021 |
| Dermatitis due to ingested food | finn-b-L12_FOODDERMAT | 199,202 | 462/198,740 | European | 16,380,428 | 2021 |

Supplimentary table 1. General information of all GWAS data.

| **SNP** | **Gene** | **Effect allele** | **Other allele** | **Chr** | **Pos** | **Beta** | **P value** |
| --- | --- | --- | --- | --- | --- | --- | --- |
| rs28473566 | DYNC2LI1 | A | G | 2 | 44012206 | -0.1752 | 8.72971e-19 |
| rs1260326 | GCKR | C | T | 2 | 27730940 | 0.0772 | 2.00798e-08 |
| rs75331444 | ABCG8 | A | G | 2 | 44069772 | 0.6995 | 3.68129e-110 |
| rs830652 | NA | A | C | 3 | 71647281 | -0.0787 | 1.86801e-08 |
| rs10935762 | TM4SF4 | T | C | 3 | 149216298 | -0.1556 | 5.00956e-14 |
| rs2290846 | LRBA | A | G | 4 | 151199080 | 0.1059 | 1.54917e-12 |
| rs17149637 | ABCB4 | A | G | 7 | 87085209 | -0.1631 | 2.0179e-14 |
| rs4738684 | NA | G | A | 8 | 59393273 | -0.1024 | 6.93745e-11 |
| rs1883016 | NEIL2/LOC124901888 | A | G | 8 | 11629637 | 0.0954 | 4.06303e-08 |
| rs686030 | TTC39B | A | C | 9 | 15304782 | 0.1241 | 2.943e-09 |
| rs2393775 | HNF1A | A | G | 12 | 121424574 | 0.0928 | 1.70687e-11 |
| rs112285002 | SULT2A1/LINC01595 | T | C | 19 | 48374320 | -0.1455 | 1.15691e-11 |
| rs570794 | FUT2/LOC105447645 | C | T | 19 | 49207651 | 0.1053 | 3.62911e-11 |
| rs6509771 | TPM3P9/ZNF761/ZNF765-ZNF761 | C | T | 19 | 53946527 | 0.0892 | 2.20298e-08 |
| rs1800961 | HNF4A | T | C | 20 | 43042364 | 0.2466 | 1.293e-09 |

Supplimentary table 2. IVs of the cholecystitis and its annotation through NCBI.

| **Exposure** | **Outcome** | **Egger_intercept** | **SE** | **Pleiotropy_pval** |
| --- | --- | --- | --- | --- |
| Cholecystitis | IL-6 | -0.0140 | 0.0077 | 0.0930 |
|  | IL-7 | -0.0042 | 0.0121 | 0.7308 |
|  | IFN-gamma | -0.0055 | 0.0080 | 0.4987 |
| **Exposure** | **Outcome** | **Method** | **Q** | **Q_pval** |
| Cholecystitis | IL-6 | Inverse variance weighted | 15.4155 | 0.5171 |
|  |  | MR Egger | 12.1284 | 0.3504 |
|  | IL-7 | Inverse variance weighted | 13.5467 | 0.4840 |
|  |  | MR Egger | 13.4192 | 0.4160 |
|  | IFN-gamma | Inverse variance weighted | 8.8584 | 0.8400 |
|  |  | MR Egger | 8.3741 | 0.8184 |

Supplimentary table 3. Sensitivity tests results of the two-step MR that the cholecystitis as exposure.

| **Exposure** | **Outcome** | **Egger_intercept** | **SE** | **Pleiotropy_pval** |
| --- | --- | --- | --- | --- |
| Allergic contact dermatitis | CCL-19 | 0.0020 | 0.0098 | 0.8392 |
|  | FGF-23 | -0.0140 | 0.0125 | 0.2718 |
|  | FIT3L | 0.0050 | 0.0089 | 0.5794 |
|  | IL-1A | 0.0226 | 0.0151 | 0.1451 |
|  | IL-18 | -0.0033 | 0.0025 | 0.1849 |
|  | IL-33 | 0.0015 | 0.0172 | 0.9309 |
|  | IL-6 | -0.0126 | 0.0186 | 0.5123 |
|  | IL-7 | 0.0398 | 0.0218 | 0.0828 |
|  | MIP-1A | -0.0005 | 0.0102 | 0.9591 |
| Exfoliative dermatitis | IFN-gamma | -0.0382 | 0.0550 | 0.4965 |
|  | IL-17A | 0.0220 | 0.0555 | 0.6942 |
|  | IL-5 | 0.0876 | 0.0524 | 0.1089 |
|  | LIF | 0.0450 | 0.0507 | 0.3839 |
| **Exposure** | **Outcome** | **Method** | **Q** | **Q_pval** |
| Allergic contact dermatitis | CCL-19 | Inverse variance weighted | 41.3911 | 0.2117 |
|  |  | MR Egger | 41.3403 | 0.1807 |
|  | FGF-23 | Inverse variance weighted | 57.4250 | 0.0038 |
|  |  | MR Egger | 32.4945 | 0.0072 |
|  | IL-1A | Inverse variance weighted | 34.0698 | 0.1640 |
|  |  | MR Egger | 53.5645 | 0.1773 |
|  | FIT3L | Inverse variance weighted | 54.1262 | 0.1409 |
|  |  | MR Egger | 53.7366 | 0.1263 |
|  | IL-18 | Inverse variance weighted | 538.4534 | 0.6409 |
|  |  | MR Egger | 536.6908 | 0.6497 |
|  | IL-33 | Inverse variance weighted | 26.1134 | 0.2468 |
|  |  | MR Egger | 26.1039 | 0.2025 |
|  | IL-6 | Inverse variance weighted | 4.5365 | 0.9717 |
|  |  | MR Egger | 4.0781 | 0.9676 |
|  | IL-7 | Inverse variance weighted | 30.9980 | 0.0737 |
|  |  | MR Egger | 26.5689 | 0.1478 |
|  | MIP-1A | Inverse variance weighted | 16.4134 | 0.6296 |
|  |  | MR Egger | 16.4106 | 0.5639 |
| Exfoliative dermatitis | IFN-gamma | Inverse variance weighted | 19.6174 | 0.3548 |
|  |  | MR Egger | 19.0756 | 0.3242 |
|  | IL-17A | Inverse variance weighted | 31.2348 | 0.5051 |
|  |  | MR Egger | 31.0769 | 0.4623 |
|  | IL-5 | Inverse variance weighted | 19.5230 | 0.6708 |
|  |  | MR Egger | 16.7377 | 0.7777 |
|  | LIF | Inverse variance weighted | 22.7689 | 0.5911 |
|  |  | MR Egger | 21.9822 | 0.5803 |

Supplimentary table 4. Sensitivity tests results of the two-step MR that the inflammatory protein as exposure.

| Mediator | β0 | β1 | β2 | Mediation effect | Proportion mediated |
| --- | --- | --- | --- | --- | --- |
| IL-6 | -0.124 | -0.079 | 0.2381 | -0.0188 | 0.1517 |
| IL-7 | -0.124 | -0.1018 | 0.2687 | -0.0274 | 0.2206 |
| IFN-gamma | -0.5555 | -0.0724 | 0.6014 | -0.0435 | 0.0784 |

Supplimentary table 5. Mediation effect caculation of the two-step MR positive results.

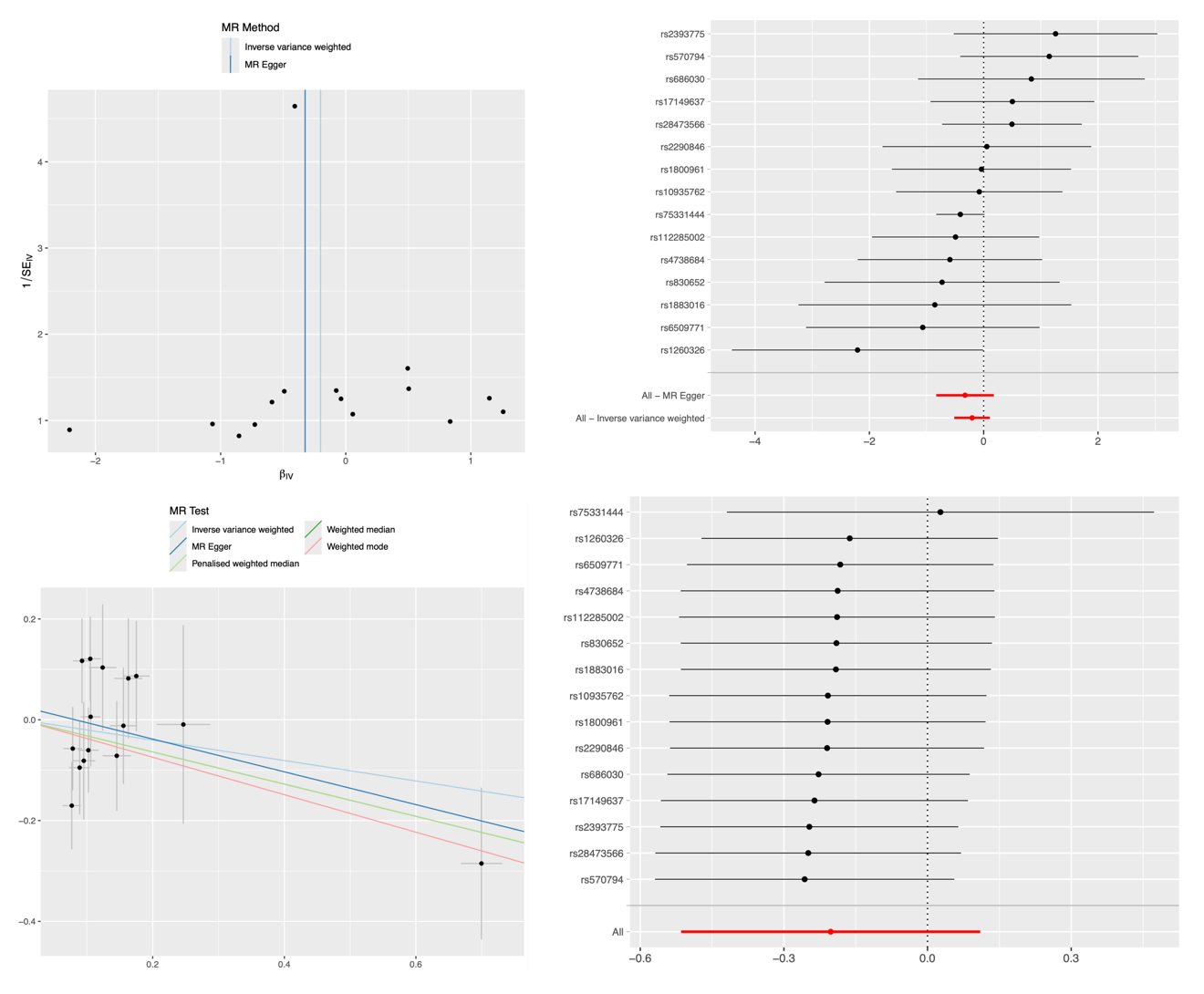

Supplimentary figure 1. Cholecystitis MR analysis on allergic contact dermatitis due to drugs in contact with skin.

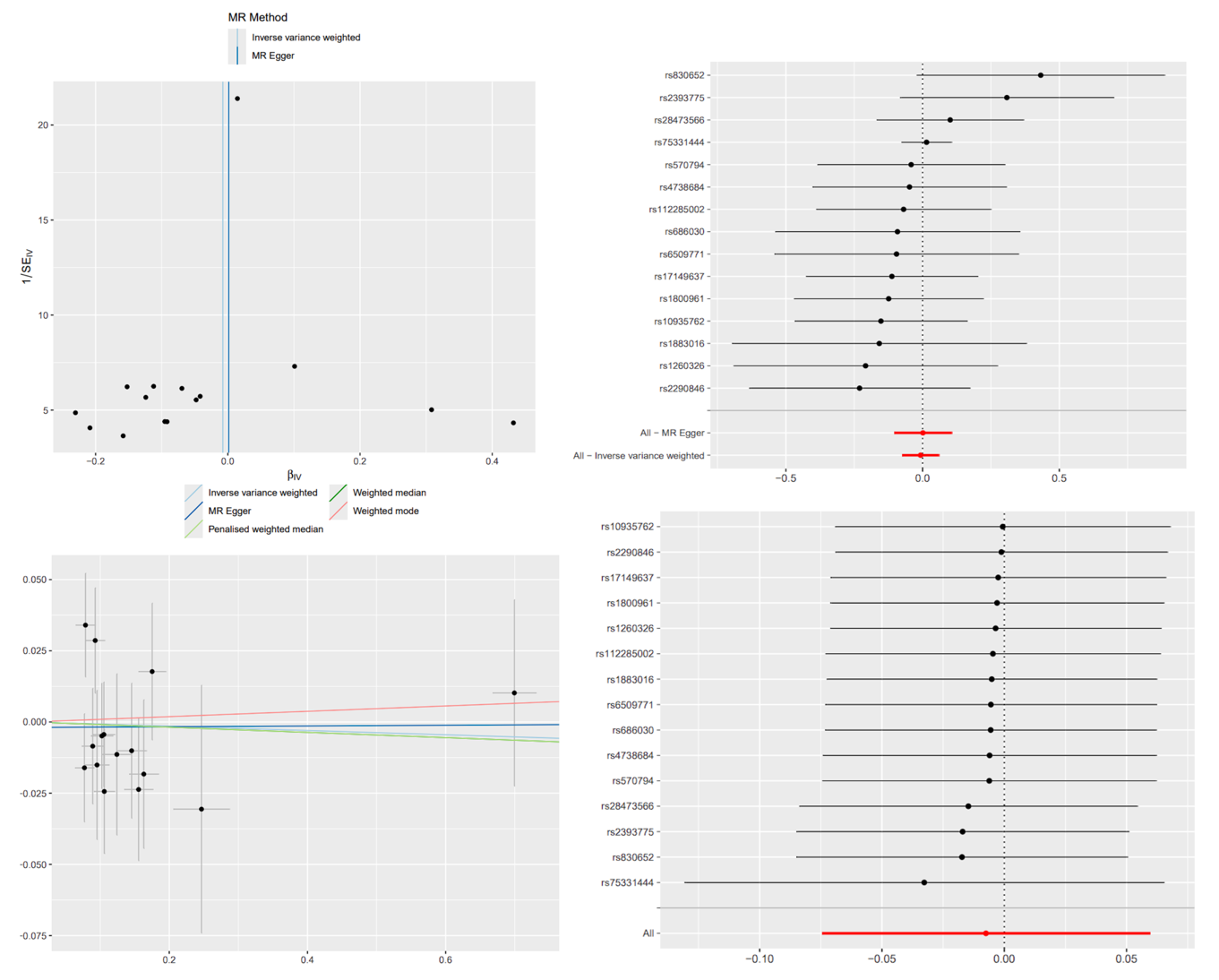

Supplimentary figure 2. Cholecystitis MR analysis on atopic dermatitis.

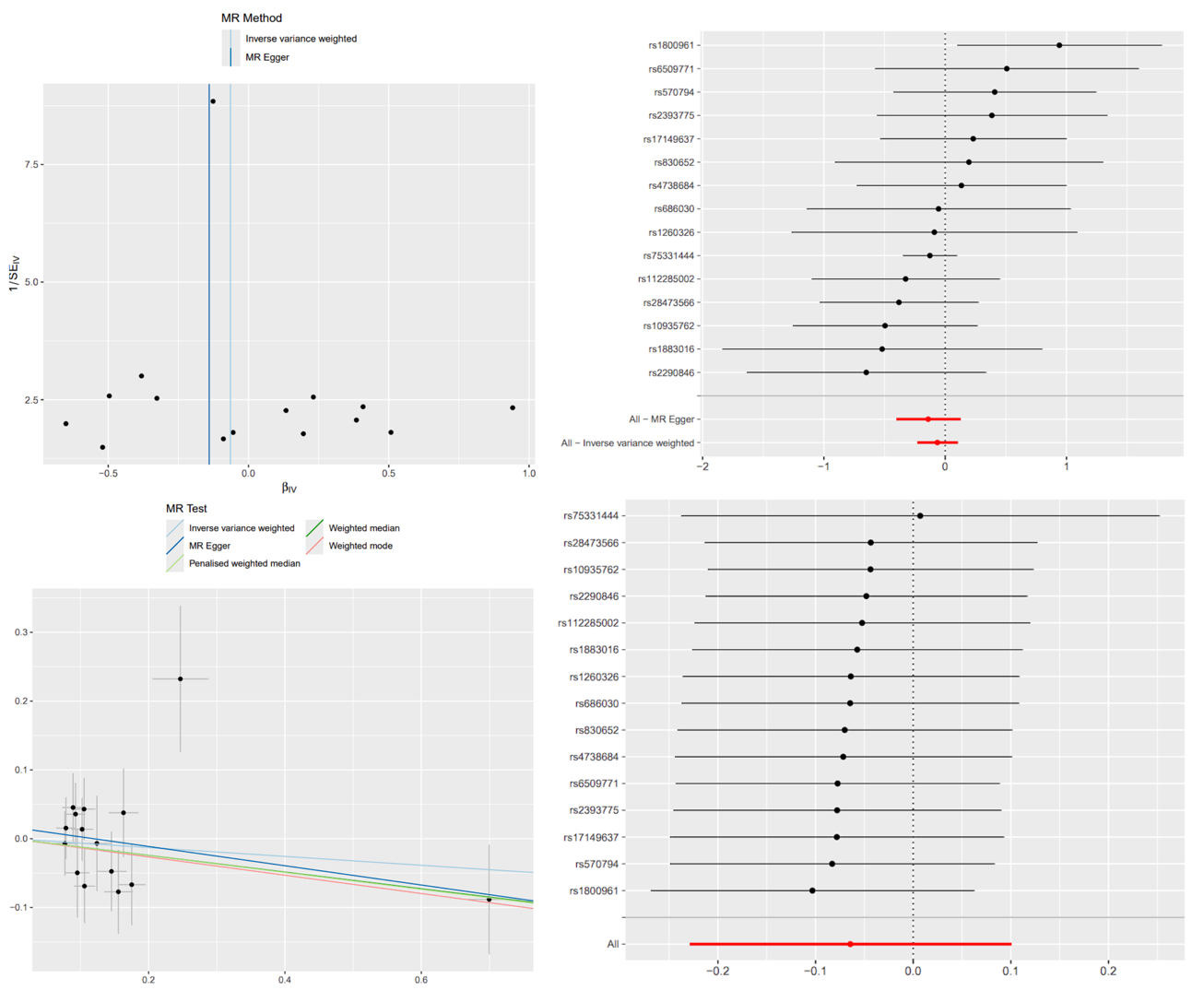

Supplimentary figure 3. Cholecystitis MR analysis on nummular dermatitis.

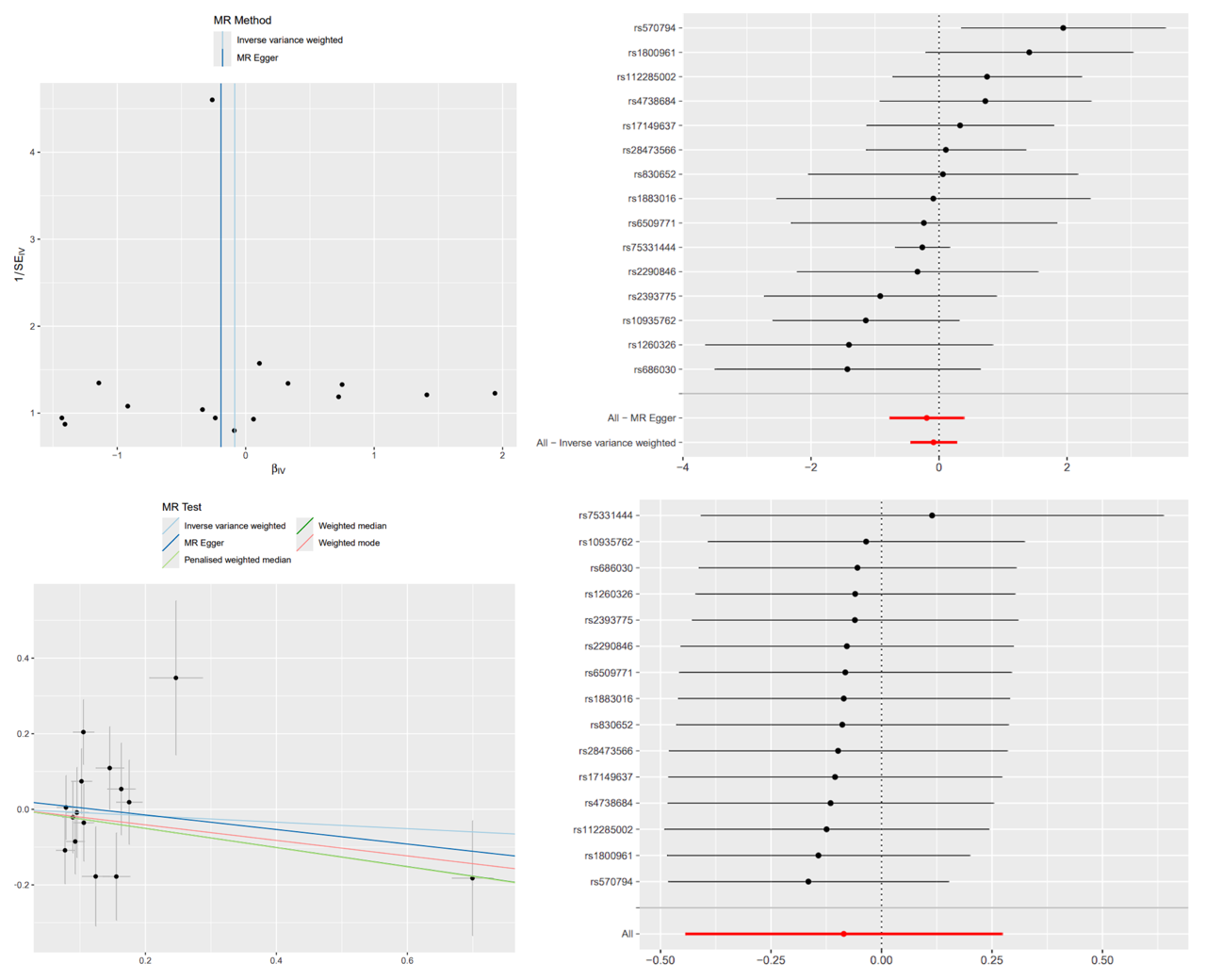

Supplimentary figure 4. Cholecystitis MR analysis on dermatitis herpetiformis.

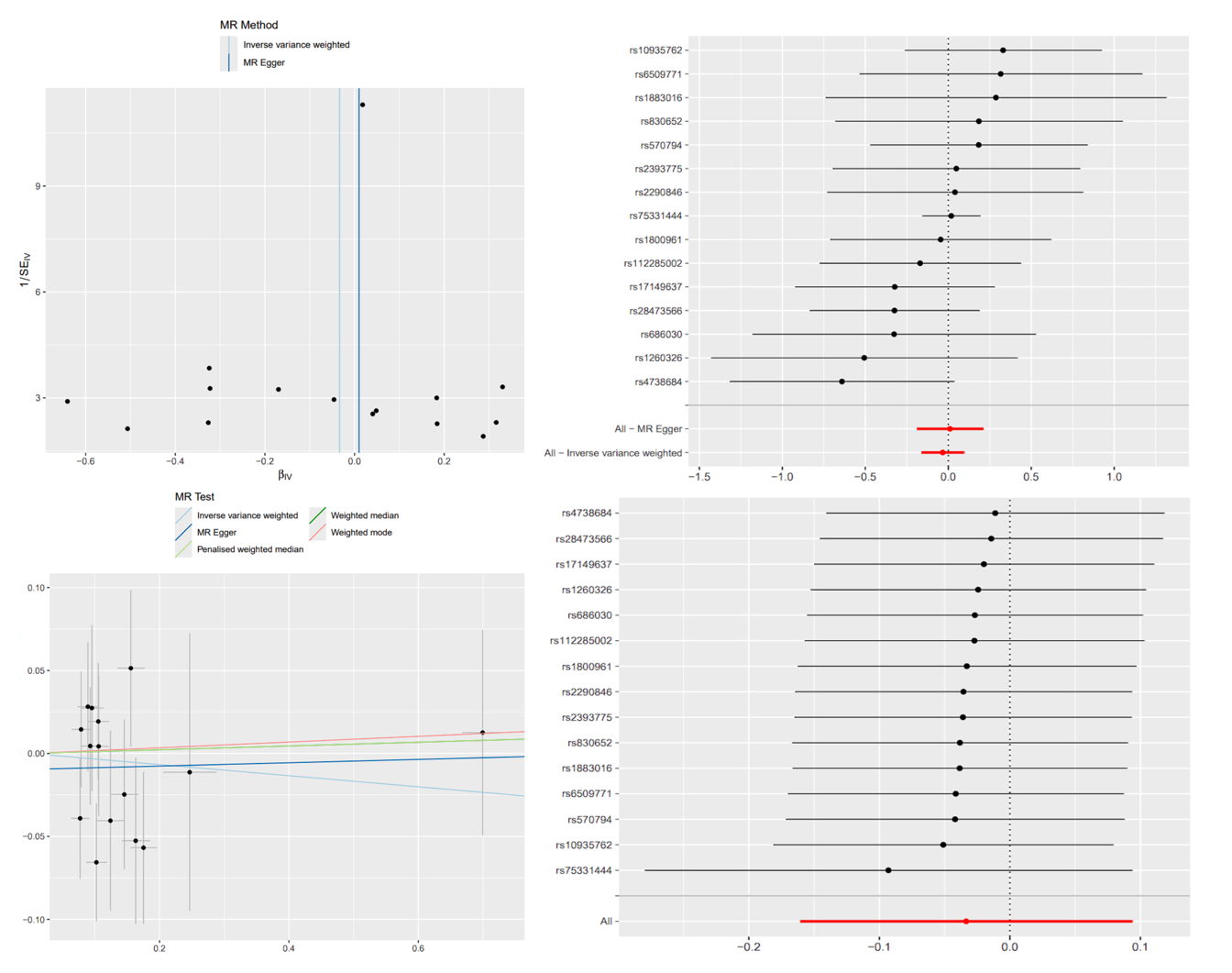

Supplimentary figure 5. Cholecystitis MR analysis on infective dermatitis.

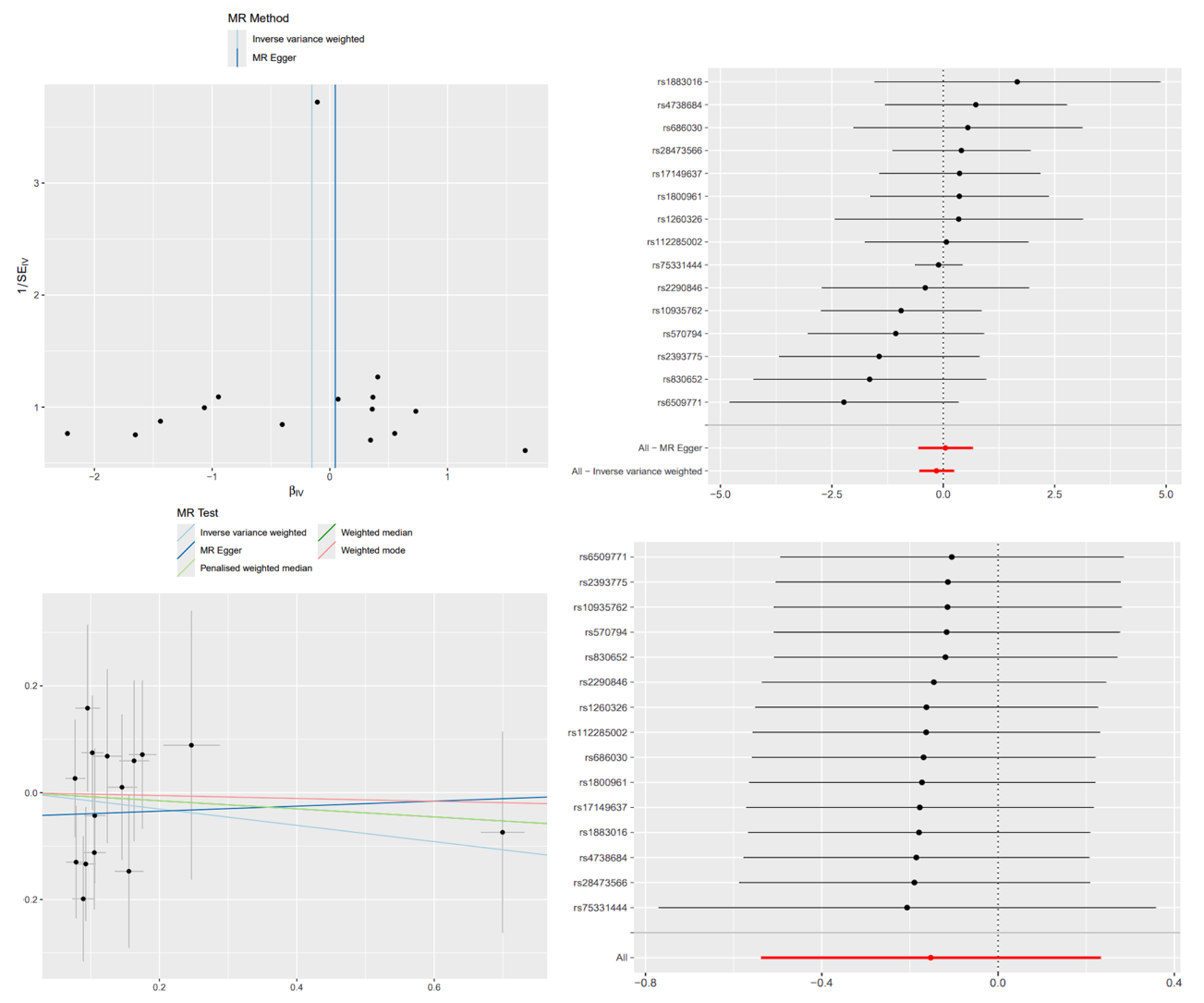

Supplimentary figure 6. Cholecystitis MR analysis on perioral dermatitis.

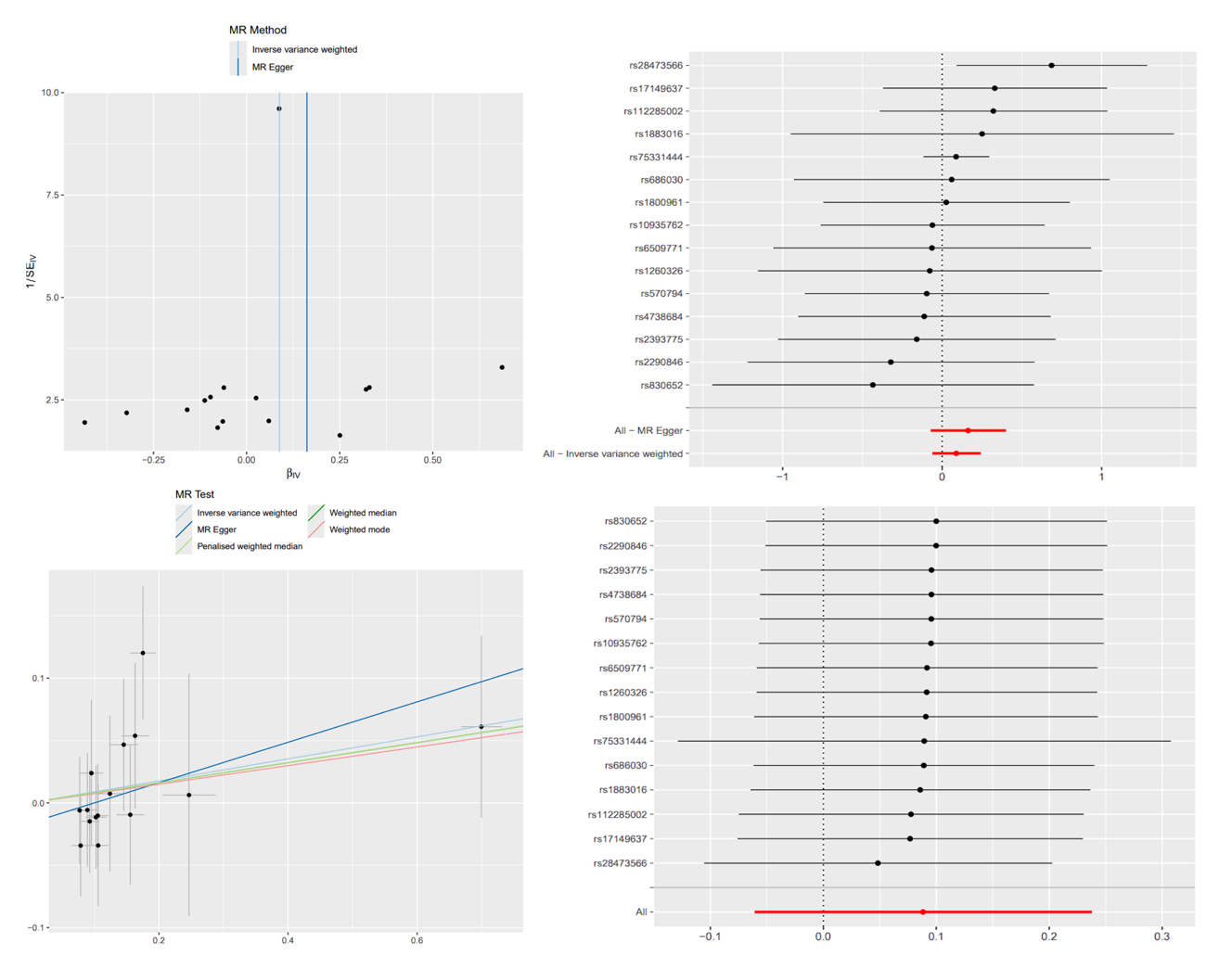

Supplimentary figure 7. Cholecystitis MR analysis on seborrhoeic dermatitis.

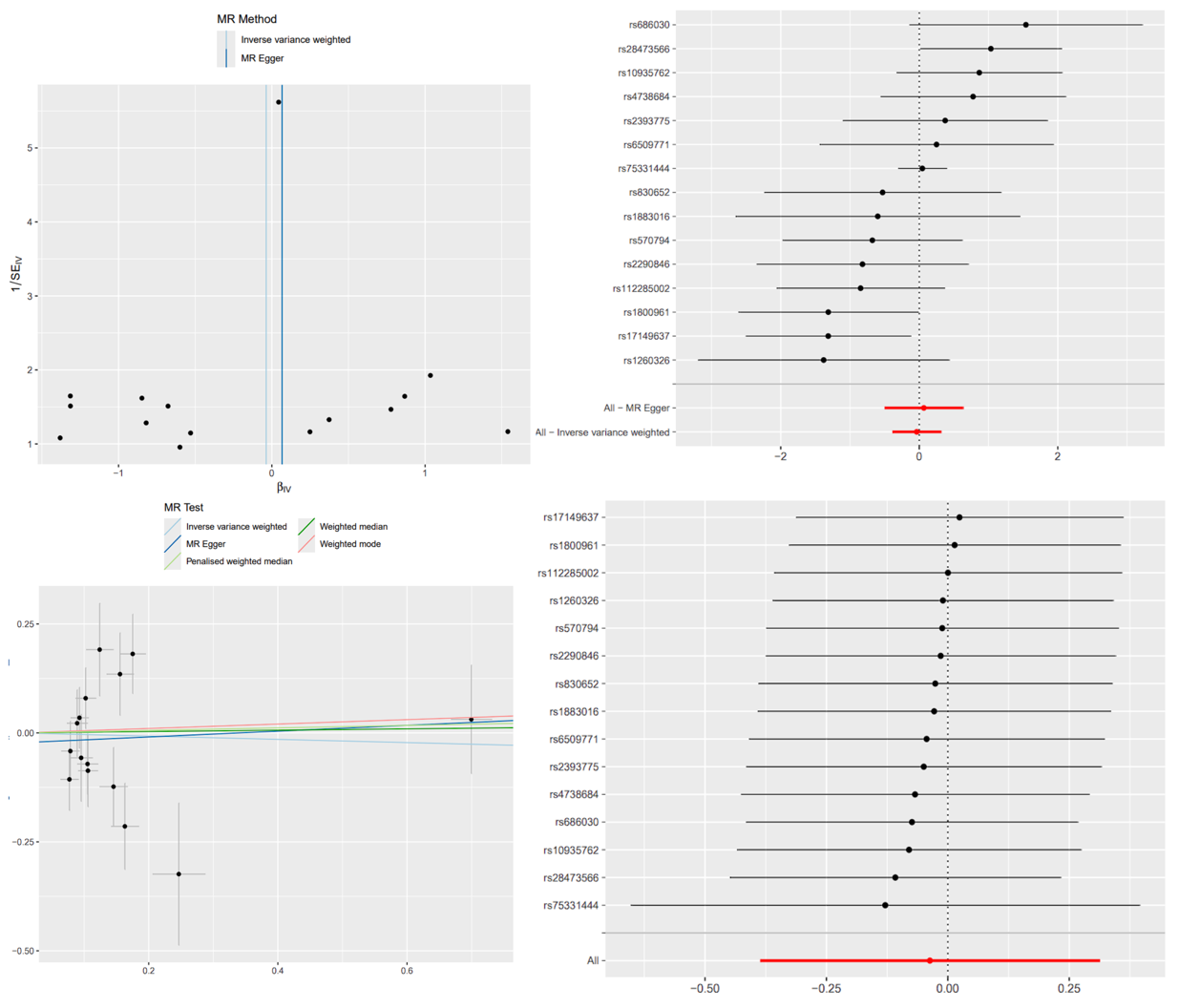

Supplimentary figure 8. Cholecystitis MR analysis on dermatitis due to ingested food.

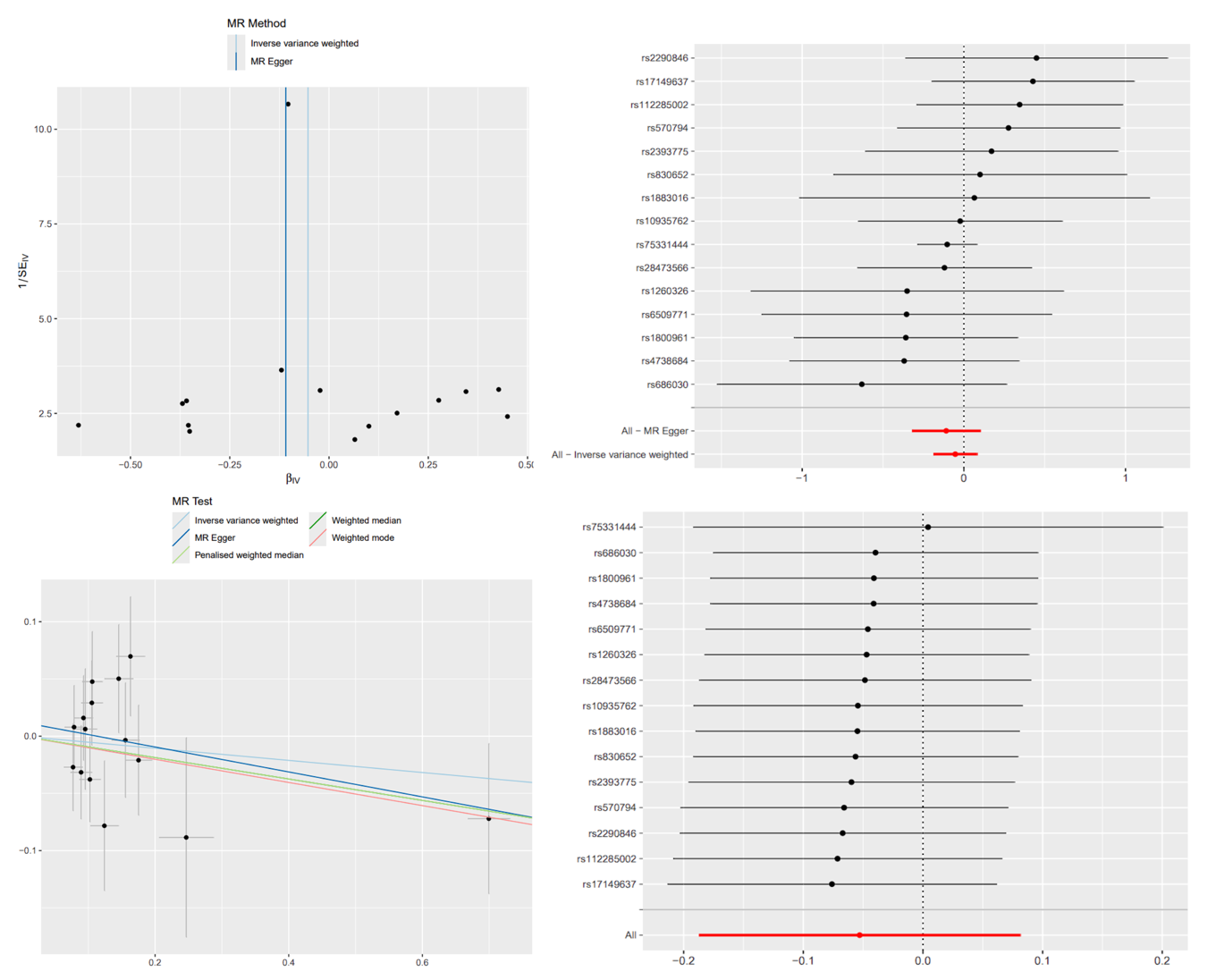

Supplimentary figure 9. Cholecystitis MR analysis on rash.
